# ‘Outsourced oxygen to the bedside’ in five countries – a qualitative implementation assessment

**DOI:** 10.64898/2026.02.20.26346703

**Authors:** Celia Blaas, Pius Mukisa, Mattias Schedwin, Hamish R Graham, Tim Baker, Ayobami A Bakare, Deepa Bishit, Elibariki Mkumbo, Jacquie Oliwa, Jacinta Nzinga, Sophie Namasopo, Máire Ruane, Aishat Adeniji, Michael Hawkes, Amarpreet Rai, Michael Njuguna, Carina King, Freddy E Kitutu

**Affiliations:** Department of Global Public Health, Karolinska Institutet, Stockholm, Sweden; Department of Pharmacy, Makerere University School of Health Sciences, Kampala, Uganda; Department of Community Health and Behavioural Sciences, Makerere University School of Public Health, Kampala, Uganda; Astrid Lindgren Children’s Hospital, Karolinska University Hospital, Stockholm, Sweden; Melbourne Children’s Global Health, MCRI, University of Melbourne, Royal Children’s Hospital, Melbourne, Australia; Department of Paediatrics, University College Hospital, Ibadan, Nigeria; Department of Emergency Medicine, Muhimbili University of Health and Allied Sciences, Dar es Salaam, Tanzania; Queen Mary University of London, United Kingdom; Clinical Research Department, London School of Hygiene and Tropical Medicine, United Kingdom; Department of Community Medicine, University College Hospital, Ibadan, Nigeria; Outline India, Gurugram, India; Department of Public Health, Institute of Tropical Medicine, Antwerp, Belgium; Health Services Unit, KEMRI-Wellcome Trust Research Programme, Nairobi, Kenya; Institute for Resilient Health Systems, Department of International Tropical Medicine, Liverpool School of Tropical Medicine, UK; Health Systems and Research Ethics, KEMRI-Wellcome Trust Research Programme, Nairobi, Kenya; Kabale Regional Referral Hospital, Kabale, Uganda; FREO2 Foundation, University of Melbourne, Melbourne, Australia; HealthPort, Lagos, Nigeria; Department of Pediatrics, University of British Columbia, Vancouver, Canada; Sanrai International, New York, USA; Center For Public Health and Development, Nairobi, Kenya; Department of Women’s and Children’s Health, International Child Health and Migration, Uppsala University, Uppsala, Sweden

**Keywords:** Oxygen, oxygen-as-a-service, qualitative research, implementation science

## Abstract

**Introduction:** Access to medical oxygen remains a challenge, with 60% of the world’s population lacking access to quality oxygen services. We explored whether ‘outsourced oxygen to the bedside’ (O2B), where private providers offer bundles of oxygen services, could be adopted and scaled across diverse health systems, to improve patient access to oxygen.

**Methods:** Qualitative interviews were conducted with healthcare workers (HCWs), facility management staff and district medical officers in India, Nigeria, Tanzania, Kenya and Uganda, who had taken part in an O2B pilot. Interviews were conducted between 19^th^ November 2024 and 25^th^ January 2025, and explored the feasibility, fidelity, acceptability, perceived benefits and weaknesses of five O2B models. Analysis used a pragmatic codebook approach, with inductive and deductive coding, informed by implementation science frameworks.

**Results:** We conducted 59 interviews, with managers and HCWs from 20 health facilities. We identified five themes relating to the potential for O2B pilots to be adopted within the health system: effect on oxygen culture, feasibility of whole of facility solutions, promotion of local ownership, being willing but unsure about the ability to pay, and the desire for hybrid tailored service models. HCWs and facility staff raised several positive factors within these themes but highlighted that understanding local facility needs and tailoring the services to embed within existing structures were important for sustainability.

**Conclusion:** Overall, the O2B service approach shows potential for implementation beyond the pilots, but more understanding of how to optimize service delivery packages to different facility needs, while also prioritizing affordability is needed.

## Introduction

Although medical oxygen is an essential component of patient care across a wide range of conditions and all age groups, 60% of the world’s population are estimated to lack access to quality, affordable oxygen services, with the largest gaps in rural, public facilities in low- and middle-income countries.^1^ Several studies have demonstrated reductions in mortality through investments in oxygen systems for health facilities with variable oxygen demand.^2–4^ However, challenges related to unreliable power supplies, infrastructure, distribution logistics and limited access to preventive maintenance remain common.^1^

This means that when oxygen equipment is available, it is not often functional.^1^ Cross-sectional facility audits of equipment availability and functionality in sub-Saharan Africa and Asia found the majority of oxygen concentrators assessed were not producing medical-grade oxygen, and oxygen cylinders are often empty, lacking regulators or equipment to enable clinical use.^1,5–8^ A study conducted in seven countries found only 24% of primary care facilities had a functional pulse oximeter,^9^ while another study from Nigeria reported only 5% of concentrators tested were functional.^5^

The 2025 Lancet Global Health Commission for Medical Oxygen Security highlighted the need for investment in biomedical engineers, better coordination of oxygen systems, sustainable financing mechanisms and regulation of oxygen markets as priority areas for innovation.^1^ While investments in medical oxygen systems increased exponentially during the COVID-19 pandemic, the majority of funds focused on equipment procurement, rather than operationalization. ‘Oxygen-as-a-Service’ (O2aaS) models have been proposed as a solution to this, where private providers offer a bundle of oxygen services, which can be tailored to health system needs. O2aaS offers can vary widely, from large-scale production and distribution to local maintenance and repair service providers. ‘Outsourced oxygen to the bedside’ (O2B) has been conceptualized as a sub-set of O2aaS, focused on providing facility or ward-level solutions that deliver the different elements needed to ensure medical oxygen availability at the point of patient need (i.e. an oxygen source, pulse oximetry, consumables, healthcare worker training, repair and maintenance).^10^

The need to consider the wider oxygen ecosystem, beyond equipment availability has been raised frequently in recent oxygen literature.^2,5,11,12^ An evaluation of a ‘hub-and-spoke’ oxygen system in Kenya, Rwanda and Ethiopia found successful implementation was linked with strong partnerships, prioritizing maintenance capacity and planning for financial sustainability (e.g. strategic pricing and local ownership).^13,14^ A qualitative follow-up of this intervention emphasized the need for strategic geographical planning to reduce transportation logistics for facilities, and the need to pair oxygen sources with consumables, pulse oximeters, infrastructure and clinical training. ^14^ Learning from different studies has underscored the need to evaluate oxygen service interventions with an implementation focus, to understand determinants and indicators of success outside of equipment alone.^15–17^

In response to this need, the Oxygen CoLab supported five pilot implementation projects, testing different O2B service packages and business models across five diverse health system settings of India, Nigeria, Uganda, Tanzania and Kenya. We aimed to explore whether O2B models could be feasibly implemented and scaled across diverse health systems to improve oxygen access.

## Methods

### Study design

We conducted a qualitative interview study using a phenomenological approach to explore the experiences of healthcare workers (HCWs), facility managers and district medical officers of O2B pilots in India, Nigeria, Tanzania, Kenya and Uganda. Interviews were conducted between 19^th^ November 2024 and 25^th^ January 2025, and explored the feasibility, fidelity, acceptability, perceived benefits and weaknesses of the O2B models. Analysis was conducted to understand implementation factors that may influence the potential for adoption and scaling of these service packages within different healthcare systems, with results presented in line with the consolidated criteria for reporting qualitative research (COREQ) checklist.^18^ This study is part of a larger cross sectional mixed-methods evaluation of the Oxygen CoLab supported O2B pilots, with the impact of O2B implementation on oxygen availability reported separately. The Oxygen CoLab programme was launched in 2020 by the ‘Better Futures CoLab’, as a collaboration between Brink and DT Global, funded by the UK Foreign, Commonwealth and Development Office. ^19^

### Framework

The evaluation of the O2B pilots was informed by two conceptual frameworks. To explore outcomes related to potential adoption and scale-up, we applied Proctor’s framework,^17^ which outlines eight domains: acceptability, adoption, appropriateness, feasibility, fidelity, implementation cost, penetration and sustainability. To explore factors that may influence or affect implementation, we used the ‘Consolidated Framework for Implementation Research’ (CFIR), developed by Damschroder et al. in 2009 and updated in 2022.^15,16^ This framework provides five domains that determine implementation - the inner setting, outer setting, implementation process, individuals and factors related to the innovation itself.

### Settings

This was a multi-country evaluation, with data collected from health facilities that had been part of the O2B pilots (**Table 1**). Details of the health systems and O2B pilots conducted in each country are provided in **Supplementary appendix 1**. The O2B pilots in each country were geographically limited to one or two states or districts, and the facilities varied in terms of clinical services offered, ownership, and pre-existing access to medical oxygen.

**Table 1:** Summary of health facilities where participants for the qualitative interviews were recruited.

| Country | State/district (urban/rural) | Ownership | Level* | Size** |
| --- | --- | --- | --- | --- |
| Nigeria | Lagos (peri-urban) | Public | Secondary | 100 |
|  | Lagos (peri-urban) | Public | Secondary | 32 |
|  | Ogun (peri-urban) | Private-for-profit | Primary | 6 |
|  | Ogun (peri-urban) | Private-for-profit | Primary | 26 |
| Tanzania | Manyara (peri-urban) | Private | Secondary | 200 |
|  | Manyara (urban) | Public | Secondary | 157 |
|  | Manyara (urban) | Private | Primary | 39 |
|  | Arusha (urban) | Public | Primary | 49 |
| India | Uttar Pradesh (urban) | Public | Secondary | 40 |
|  | Uttar Pradesh (rural) | Public | Secondary | 35 |
|  | Uttar Pradesh (urban) | Public | Secondary | 10 |
|  | Uttar Pradesh (rural) | Public | Secondary | 30 |
| Kenya | Nairobi County (peri-urban) | Public | Primary | 22 |
|  | Nairobi County (peri-urban) | Private | Primary | 14 |
|  | Nairobi County (peri-urban) | Faith based | Primary | 35 |
| Uganda | Kanungu (rural) | Private-not-for-profit | Primary | 6 |
|  | Kanungu (rural) | Private-not-for-profit | Primary | 36 |
|  | Kanungu (peri-urban) | Private-for-profit | Secondary | 27 |
|  | Kanungu (rural) | Public | Secondary | 100 |
|  | Kanungu (rural) | Private-not-for-profit | Secondary | 155 |

\*Facility levels were classified as primary (level 1), secondary (level 2) or tertiary (level 3), based on the Lancet Global Health Commission on Medical Oxygen Security and WHO Emergency and Essential Surgical Care definitions.<sup>1,20</sup>
\*\*Number of inpatient beds

### Recruitment

Health facilities were purposively selected from the list of facilities where O2B pilots were implemented, through discussion with the O2B implementation providers. We took a maximum variation approach, selecting facilities that represented different sizes, geographies, levels of care, time implementing the O2B model, and pre-existing oxygen equipment availability. In each country, we selected a minimum of three facilities for qualitative data collection (**Table 1**). Interview participants were recruited from the facilities, using purposive sampling for facility managers and convenience sampling for HCWs, with the aim of conducting a minimum of two interviews at each facility. Where possible, we included participants from a range of cadres, ages, genders and levels of experience.

The facility managers at all facilities were approached for recruitment, with a nominated in-charge interviewed in instances where the manager was unavailable. HCWs on duty at the time of data collection were approached by the qualitative researcher. In some facilities, additional interviews were conducted when differing perspectives emerged amongst HCWs, to ensure data saturation was achieved.

### Data collection

Interviews were conducted by experienced qualitative researchers in each country (AAB, JN, DB, PM, EM). Researchers were a mix of female and male genders, with doctoral or master’s level degrees in health and social sciences. None of the researchers were related or known to study participants, and none were affiliated with the O2B implementers. Interviews were conducted face-to-face in locations convenient to participants, including offices at health institutions or their facility, with one instance of a phone interview being conducted. No repeat interviews were done.

Eligible participants were provided with written study information and were given the opportunity to ask questions before the researcher asked for informed consent. Interviews were either conducted in English or the local language (Kiswahili, Yoruba and Hindi), dependent on participant preference. Interviews were audio-recorded with participant consent, and then transcribed and translated into English where necessary, by the researchers who conducted the interviews. Transcripts were not returned to participants for clarification. In instances where participants declined to be audio recorded, written notes were taken.

Interviews used a semi-structured interview guide (**Supplementary appendix 2**). An overall guide was developed by CB, PM, MS, FK and CK, and was adapted to the different settings by the local researchers (AAB, JO, JN, DB, PM, EM) following an online training which presented the project aim, objectives and O2B implementation pilot. After the first facility visit in each country, CB and the local qualitative researcher communicated about clarifications needed, rewording of questions, and recruitment progress and strategy.

### Data analysis

Interviews were analysed using a pragmatic framework approach, with a combination of inductive coding, and deductive coding based on predefined themes from the CFIR and Proctor frameworks.^16,17,21^ The codebook was developed by CB and reviewed by PM, CK and FK, then updated by CB and PM using a flexible approach to include new themes and ideas that emerged during coding. After familiarization with the transcripts, seven of the interviews were double coded by CB and PM to promote consistency throughout the analysis, and inter-coder reliability. The remaining interviews were then coded by either CB and PM, with any uncertainties discussed with CK. Codes were discussed, refined and organised into themes by CB, which were presented to the local qualitative researchers for input and feedback before finalization. Analysis was done using NVIVO Version 15.0 (QSR International, Doncaster, Australia).

### Ethics

All participants provided informed written consent for interviews, and audio recording. Ethical approvals were obtained from ethical review boards in each country: Monk Prayogshala Institutional Review Board, India (reference: #156-024); Maseno University Scientific and Ethics Review Committee, Kenya (reference: MSU/DRPI/MUSERC/01361/23); National Commission for Science, Technology and Innovation, National Commission for Science, Technology and Innovation, Kenya (license number: NACOSTI/P/24/37427, reference: #673262); Health Research and Ethics Committee, LASUTH, Lagos State, Nigeria (reference: LREC/06/10/2498); Health Research Ethics Committee, Ogun State, Nigeria (reference: OGHREC/467/2024/290/APP); National Institute for Medical Research, Tanzania (reference: NIMR/HQ/R.8a/Vol.IX/4743); Ugandan National Council for Science and Technology, Uganda (reference: HS5437ES); Makerere University School of Health Sciences REC, Uganda(reference: MAKSHSREC-2023-600). Approval was granted by the Swedish Ethics Review Authority for the processing of personal data (reference: DNR 2024-00868-01).

### Patient and public involvement

Patients and the public were not involved in the design or execution of the study.

## Results

We conducted a total of 59 interviews across 20 health facilities (**Table 2**). No participants declined to be interviewed. We identified five themes relating to the potential for O2B pilots to be scaled within the health system: effect on oxygen culture, feasibility of whole of facility solutions, promotion of local ownership, being willing but unsure about the ability to pay for the service, and the desire for tailored service models (**Supplementary appendix 3**).

**Table 2:** Summary of participant characteristics.

|  | Nigeria | India* | Tanzania | Kenya | Uganda |
| --- | --- | --- | --- | --- | --- |
| **Total** | 16 | 11 | 12 | 9 | 11 |
| **Cadre** |  |  |  |  |  |
| District health officer | - | 1 | 1 | - | - |
| Facility in-charge | 1 | 3 | 2 | 3 | 4 |
| Doctor | 4 | 2 | 2 | - | 1 |
| Nurse / midwife | 3 | - | 5 | 4 | 4 |
| Other clinical staff** | 2 | - | - | 1 | - |
| Pharmacist | - | 5 | - | - | 1 |
| Biomedical / technical staff | 1 | - | 1 | 1 | 1 |
| Administrative staff | 5 | - | 1 | - | - |
| **Gender** |  |  |  |  |  |
| Male | 5 |  | 6 | 4 | 10 |
| Female | 11 |  | 6 | 5 | 1 |

\*Gender was not collected in India.
\*\*In Nigeria Community Health Officers (CHOs) and Community Health Extension Workers (CHEWs) also provide facility-based services.

### Theme 1: O2B models were perceived to improve the oxygen culture and oxygen service readiness

Participants from all countries, except India, consistently described a perceived added value of the O2B pilots on oxygen culture, resulting in increased HCW’s trust in their facilities’ readiness for oxygen provision. Staff confidence in oxygen readiness was attributed to better quality, easier to use oxygen equipment from the O2B providers, including sufficient equipment to treat multiple patients simultaneously. In particular O2B staff being readily available to refill, replace or fix equipment when needed was appreciated. Participants perceived the O2B providers to be reliable and often reported they were filling a service gap. In India however, while the O2B solution was positively received, it was less appreciated given other oxygen interventions were already in place in many facilities.

HCWs described the knock-on impact of reliable oxygen access on the use of pulse oximetry and initiating oxygen treatment practices. Interviewees described an increase in pulse oximetry measurements done, including for patients that may not look critically unwell, and several stated that nurses felt more empowered to start oxygen for hypoxaemic patients.

> *“Before, we were not so much concentrating on the percentage of the oxygen levels of a child, but nowadays we are” Kenya, nurse*
>
> *“They have built our capacity in many things, especially for young children. So, all my employees are aware about oxygen. For example, even if a nurse receives a patient, they cannot wait for the doctor, you will find that they have already provided [pulse oximetry and oxygen] services because they are all aware” Tanzania, facility in-charge*

### Theme 2: providing a holistic package of oxygen services was highly desired

A unique and important feature of the O2B models described was the packaging of oxygen services, however some interviewees expressed doubt about service providers being able to meet all of the facilities oxygen needs. The extent to which each of the O2B providers included these services (i.e. training, maintenance and repair, consumables) varied between and within countries (**Supplementary appendix 1**), however interviewees were unanimously positive about the packaging of services. For instance, facility managers highlighted the convenience and reduced logistics related to oxygen procurement and supply.

> *“Apart from the oxygen, you pay for the service, the cylinder supply, the transportation*… *to me, carrying the oxygen up and down. I prefer to pay for the service like that. Let them bring it*… *it gives you [peace] of mind”. Nigeria, finance officer*

Where service elements were not included in the O2B package, these were consistently requested by interviewees, reiterating the perceived value of a comprehensive service package. Pulse oximeters and oxygen consumables were the most desired add-ons, and particularly paediatric-specific delivery equipment that facilities do not always have.

> *“While pulse oximeters have been provided, an increase in their number would be helpful” India, pharmacist*
>
> *“It would be better if they gave to us all the equipment used in oxygen delivery to the patient, like the pulse oximeter, oxygen masks” Uganda, nurse*

When asked about additional desired services, participants from all countries, except India, asked for more oxygen equipment (implying that they still had unmet oxygen needs). This included requests to expand O2B services to other wards in facilities, when the pilot was delivered as a ward-based solution like in Tanzania:

> *“I mean, this [O2B] system has gone to specific beds. But if we got concentrators, they could be placed even in other wards, making them movable” Tanzania, procurement officer*

Backup oxygen sources, and additional production capacity were also often requested, including mobile oxygen concentrators and cylinders. In Nigeria specifically, several respondents noted the ‘O2 cube’ production capacity was insufficient to meet facility oxygen demands, resulting in the O2B provider shifting to source additional cylinders from an external provider.

Finally, participants expressed concerns that O2B providers were unable to address facility infrastructural challenges – particularly power supply and transportation of oxygen equipment between wards - which ultimately limited oxygen access for patients. Participants consistently asked for back-up power to be included, with suggestions including an oxygen-specific generator, or additional solar power with batteries to run equipment uninterrupted. Many interviewees also expressed a desire for piped oxygen to the bedside to avoid the hassle of moving cylinders between beds.

> *“I think when we have piped oxygen is better, there won’t be system of carrying cylinders around and power interruptions” Uganda, nurse*

### Theme 3: prioritizing local ownership and awareness of O2B services could improve longer term sustainability

According to interviewees, an important part of building local ownership was building on-site knowledge for monitoring, maintenance and repair of oxygen equipment to promote long-term use and longevity of equipment. Several participants reported a reliance on O2B staff to monitor equipment functionality, which allowed facility staff to focus on other things, but there were also examples given of broken equipment only being identified when O2B providers conducted regular monitoring. This meant there were risks of interruptions to oxygen as facility staff were unable to solve problems independently. The importance of collaboration and strong working relationships was consistently highlighted to improve functionality and workflow of the O2B pilot models.

> *“For oxygen, we totally rely on Health Port to sort themselves out on that, so we don’t really, there’s no interference, the biomedical engineer focuses on instruments, equipment and all that’s not oxygen” Nigeria, doctor*
>
> *“So that the time the thing is malfunctioning, and you have a child on oxygen, so you are thinking who do I call? Because if we say you shout for help no one will come to help you, what will they help you with? That is where the problem is. You are unable to find the technician” Tanzania, nurse*

Linked to this was also the request for clinical and equipment training to be included to support longer term sustainability of the O2B models, as well as repeat or refresher training alongside recurrent sessions to educate new staff and re-enforce practice.

> *“Currently staff are not adequately trained on how to use oxygen concentrators or determine the correct oxygen dosage for children. Providing such training would be highly beneficial, as many staff members lack this essential knowledge” India, facility in-charge*

Participants stressed that embedding O2B services within facilities relied on collaboration between facility and O2B staff, awareness of the model, and local buy-in. When participants described issues with integration, this was attributed to a lack of familiarity with the O2B pilots amongst facility staff. For example, not realizing that maintenance of equipment was included or not knowing how to operate their equipment, often amongst new staff or those working in ward areas without access to the intervention. In the case of FREO2’s Oxylink concentrator system in Tanzania, one nurse reported that not all clinical staff were aware of the automatic switch over to back-up cylinders during power outages. Several participants made suggestions to improve awareness and local ownership through local mentors or “oxygen champions” who could motivate staff to use equipment and be the contact person if issues arose.

> *“I just emphasize, there should be a champion of that system. Even if they stop the service so that the person continues or if there is a challenge, they can solve it. Because there will be a disconnect” Tanzania, nurse*
>
> *“Sustainability is about if we take ownership” Nigeria, doctor*

### Theme 4: despite a willingness to pay for O2B packages, the feasibility of paying was uncertain

Willingness to pay for O2B service packages was positively influenced by oxygen being considered a high priority, with participants stating oxygen is an essential part of critical care. Additionally, the experience of facing challenges in oxygen procurement, particularly the responsibility for facilities to procure this alone, contributed to facility managers expressing a willingness to pay for the O2B service package. Participants expressed their desire not to return to their previous method of oxygen procurement, and they consistently expressed a dedication and willingness to find payment mechanisms.

“*We are required to strive to find any way possible for this system to continue to stay” Tanzania, nurse*

However, some pilots were offered free to facilities, and a ‘donor mindset’ was seen in some respondents where they were surprised at being asked about payment for the O2B services following the pilot phase.

Cost and affordability were raised as issues by some respondents, and different perspectives were seen. For example, in Nigeria one respondent stated that the cost of O2B services were too high, while another felt the higher cost was justified and represented good value for money given the reliability, convenience and reduced logistics.

> *“It [the O2B service] is not cost effective […] the amount of cost we spend nowadays is far, far more than when we were using external vendor” Nigeria, accountant*
>
> *“Apart from the oxygen, you pay for the service, the cylinder supply, the transportation […] even the transportation is more than that. If you want to transport oxygen, you know how heavy oxygen is. Even some vehicles will not want to carry it. I: So, you find [the O2B service] cheap for you? P: Yes, I prefer that” Nigeria, biomedical engineer*

Context-specific payment mechanisms were stated to strongly influence the facilities ability to transition to a paid O2B model. In the pilots where patients pay for oxygen, some participants stated that costs are passed onto patients, and so higher costs would be passed on those who cannot afford higher prices. Participants from Nigeria specifically appreciated the flexibility of payment plans, including alternative schemes for payment when patients were unable to pay, and maintaining supply in instances of delayed payment.

> *“We’re not charging patients more but what I’m saying is that it increases our own cost because [O2B provider] price is higher than what we use” Nigeria, doctor*

Additionally, participants indicated that mechanisms to meter and record patient use of oxygen did not always align with how O2B providers charged for oxygen consumption. Several interviewees commented that charging patients for a small volume of oxygen is difficult (e.g. for stabilization), but if O2B providers have metered the use, facilities are required to cover the cost. In other instances where patients do not pay for oxygen and facility budgets are government allocated, participants stated that facilities would struggle to pay higher costs for O2B services given the lack of oxygen-specific budgets, small facility revenues, or lack of additional government funds.

> *“It will be difficult, especially in terms of maintenance, it will be a challenge. Because the level of this centre its internal revenue is very small compared to supply […] otherwise, if we had that ability we would have bought them before. We got that is just donation, but if this donation has a cut point it means the centre will go back there.” Tanzania, facility in-charge*

### Theme 5: mixed oxygen sources were desired, and should be integrated with other providers and existing infrastructure

Participants stated the need for multiple sources of oxygen within a facility to bridge gaps and ensure consistent, reliable availability of oxygen for patients. While some O2B service packages included back-up oxygen sources, the single source of oxygen provided by others was seen as a limitation. Participants raised the need for concentrators in wards where cylinders could not be transported, or inversely, the need for backup cylinders to cover for surges in oxygen demand, critically unwell patients or power outages. Interviewees asked for hybrid solutions, and emphasized this through conflicting and contrasting opinions regarding the best source of oxygen for the facility. For instance, while some expressed a desire for cylinders due to their higher oxygen flow rates, others preferred concentrators given their ease of use.

> *“Initially, I requested concentrators. But now, I think if the concentrators are not enough, we could use alternate oxygen cylinders” India, facility in-charge*

Many participants stated there was no preferred alternative to the O2B providers, although they did not necessarily stop procuring from other vendors simultaneously. While some competitors included equipment maintenance or offered transportation services for cylinders, these were not regarded as a ‘package of services’ in the way the O2B pilots were. However, some underscored the need for O2B providers to integrate and work synchronously with other existing suppliers to meet the full oxygen needs of the facility.

> *“Yes, we patronize the other vendor because of the big size of their cylinder”. Nigeria, biomedical engineer*

## Discussion

We assessed five different O2B pilots, to explore if this outsourced service model could be routinely adopted within diverse health systems, and what lessons are important to support implementation. We found that HCWs and facility managers raised several positive factors, and understanding local facility needs in order to tailor and embed services within existing structures will be important for effective adoption and scale up – the precursors for sustainability planning. We observed some consistent experiences across the settings, including the positive impact on oxygen culture, requests to include back-up electricity supply and address infrastructure gaps. However, willingness and ability to pay for the service model and perceived ability of O2B providers to meet all the oxygen needs of facilities were more varied.

An interesting tension was seen in the data, with HCWs often requesting further clinical and equipment training from the O2B providers so they could take more responsibility for maintenance and repair. However, concurrently, having an external provider that was responsible for maintenance and repair was seen as a key value-add of the O2B package. This may reflect the varied services offerings between the O2B providers, but also high clinical staff change-over can result in new staff that are unfamiliar with the O2B equipment and on-going pilots. This can increase the potential for broken equipment or prolonged down-times if they are unaware that they could call for external repairs and replacements. Whilst it is unrealistic for O2B providers to be solely responsible for training facility staff on oxygen and equipment use, inadequate training could affect the cost-effectiveness of the O2B intervention. The suggestion of ‘oxygen champions’ was given as a solution to this, with designated facility staff that serve as a point of call for facility staff to inform and train staff on appropriate use of the equipment and promote in-service culture. This approach has been demonstrated to have a positive impact in other contexts, with facility leadership a core aspect of promoting quality of care. ^22^

The potential additional impact of the O2B intervention on clinical practice culture and broader oxygen care in facilities where they are implemented should be appreciated. A positive feedback loop exists between user’s trust in an intervention, and sustained uptake and use. Studies have also shown that when staff witness the benefits of effective oxygen use firsthand, they are more likely to adopt changes in their practice, including increased use of pulse oximetry and oxygen equipment for patients that need it.^12^ A study in Nigeria exploring adoption of pulse oximetry practices found strategies that boost its attractiveness to users work through the perceived *relative advantage* for the user, including identifying hypoxemia and guiding oxygen therapy. ^22^ We found the O2B intervention appeared to increase HCW confidence to start oxygen treatment, with improved trust in their facility’s ability to manage hypoxaemic patients. This then acted as a driver of increased oxygen demand – both from HCWs and patients. A positive feedback loop of improved quality of care, leading to improved care-seeking and treatment acceptance, and therefore earlier and more effective treatment can emerge. ^23^ For O2B pilots, this has an implication for potential scale, if their service increases oxygen demand within facilities.

Trust in the O2B models also positively impacted use and demand through staff perceiving the O2B models as improving the quality of oxygen equipment and noted that equipment was easy to use. Current literature demonstrates successful uptake of oxygen interventions is affected by ease of use, including quality, user-friendly equipment and perceived acceptability. ^12,24,25^ This supports the claim that improving oxygen *use* in facilities through interventions is equally as important as improving oxygen supply,^1,25,26^ and factors such as the inner setting, acceptability and penetration are linked to successful implementation. ^16,17^

The two areas where more reflection is needed were the financing approaches for the service models, and the desire for whole-of-facility oxygen interventions that can integrate different oxygen sources. On the latter, requests from interviewees in facilities where the O2B pilot was ward specific (e.g. FREO2’s paediatric and neonatal focus) consistently included expanding O2B to all wards in the facility, and more generally participants requested more than one type of oxygen source (i.e. providing concentrators when cylinders were in the model, or vice-versa). While the request to expand the O2B service can be seen as a positive, it also highlights that a “one size fits all” approach may not always be appropriate. Often facilities operate with multiple modes of oxygen delivery simultaneously,^1^ which historically have resulted in complex and complicated procurement, management and maintenance. ^27^ However, HCWs reinforced the perceived importance of including different forms of equipment in the model as back-ups and to provide flexibility in deployment. Therefore, O2B models that are flexible and can adapt their equipment offering and integrate with existing providers to meet holistic facility needs may be more viable.

Financial models that ensure the implementation costs for O2B service delivery can be recovered are critical to sustainable and routine adoption beyond these pilots. While our results demonstrated oxygen is a priority and facilities expressed a willingness to pay for O2B service packages, their capacity to pay was less clear in some cases. Mechanisms to transition from fully or partially subsidized models were not frequently discussed by participants. In Nigeria and Uganda, facilities paid for the services, and in Nigeria while facility spend on oxygen increased, several respondents felt it represented value for money. However, in contexts where oxygen is paid for out-of-pocket by patients, increased oxygen costs run the risk of being passed onto patients, posing an issue of equity. Given high out-of-pocket costs have already been widely reported as a barrier to care, ^23,25,28,29^ an equity perspective needs to be considered when developing O2B financing models. In settings where facility budgets are government allocated, regional or ministry-level agreements rather than facility level agreements will likely be needed, and could support lower costs through economy of scale. Future research on context-specific economic models to balance costs and volumes of oxygen demand within facilities, and the cost-efficiency of other value-add services such as oxygen training, maintenance or infrastructure is needed.

We had three key limitations. Firstly, we chose to take a purposive approach to sampling facilities and participants, through consultation with the O2B providers and HCWs present on site at the time of data collection. This was done to try and ensure a range of facility types, sizes and geographical locations, however, this does not necessarily mean we captured the full range of experiences with the O2B pilots. Utilization of a standardized interview guide may also mean nuanced experiences between different HCWs may have been missed. Second, social desirability bias may have affected the participant’s responses and over-represented the potential positive benefit of the O2B intervention or resulted in perceptions that were not voiced during the interviews. The qualitative researchers were independent from the O2B providers, and this was explained to participants to try and mitigate this. Finally, the analysis approach focused on commonalities and differences between the pilots and therefore may have missed *within* country variations, and underlying mechanisms driving the differences.

Overall, the O2B pilots’ approach to service package implementation show potential for routine adoption, with successful integration within existing facility oxygen systems having potential for long term impacts on quality of care. However, uptake within the broader health system and the capacity for health facilities to pay is highly context dependent and should consider equity in contexts where oxygen is paid for out-of-pocket by patients. Therefore, more understanding of how to optimize service delivery packages to different facility and health system needs, whilst being able to achieve an economy of scale that results in a viable business model is needed.

## Authors contributions

The study was conceived by CK, FEK, MS, PM, CB, HRG and TB, with funding acquisition by CK and FEK. The O2B pilots were led by MN, MH, MR, AA and AR. Protocol and tool development was led by MS, CB and PM, with support from CK and FEK. Data collection was conducted by DB, AAB, JN, JO, EM and PM. Analysis was conducted by CB and PM, with support from CK and FEK. The manuscript was drafted by CB, with support from CK. All authors read and contributed to the final manuscript.

## Acknowledgements

We would like to thank all the healthcare workers and facilities for giving us their time and data and allowing us to discuss their experiences. We would also like to acknowledge the data collectors for their contributions.

## Data availability

Given the qualitative nature of the data, transcripts cannot be sufficiently anonymised for Open Data Access. Data can be requested for the purposes of further academic research, by contacting Carina King and Freddy Eric Kitutu. Requests will be reviewed and discussed with the site qualitative leads, and approval from local ethical committees will need to be sought.

## References

1. Graham HR, King C, Rahman AE, Kitutu FE, Greenslade L, Aqeel M, et al. Reducing global inequities in medical oxygen access: the Lancet Global Health Commission on medical oxygen security. Lancet Glob Health. 2025 Mar;13(3):e528–84.

2. Duke T, Wandi F, Jonathan M, Matai S, Kaupa M, Saavu M, et al. Improved oxygen systems for childhood pneumonia: a multihospital effectiveness study in Papua New Guinea. Lancet Lond Engl. 2008 Oct 11;372(9646):1328–33.

3. Graham HR, Bakare AA, Ayede AI, Eleyinmi J, Olatunde O, Bakare OR, et al. Cost-effectiveness and sustainability of improved hospital oxygen systems in Nigeria. BMJ Glob Health [Internet]. 2022 Aug 10 [cited 2025 May 6];7(8). Available from: https://gh.bmj.com/content/7/8/e009278

4. Fashanu C, Mekonnen T, Amedu J, Onwundiwe N, Adebiyi A, Omokere O, et al. Improved oxygen systems at hospitals in three Nigerian states: An implementation research study. Pediatr Pulmonol. 2020 Jun;55 Suppl 1:S65–77.

5. Bakare AA, Graham H, Ayede AI, Peel D, Olatinwo O, Oyewole OB, et al. Providing oxygen to children and newborns: a multi-faceted technical and clinical assessment of oxygen access and oxygen use in secondary-level hospitals in southwest Nigeria. Int Health. 2020 Jan 1;12(1):60–8.

6. Graham HR, Olojede OE, Bakare AA, Iuliano A, Olatunde O, Isah A, et al. Measuring oxygen access: lessons from health facility assessments in Lagos, Nigeria. BMJ Glob Health. 2021 Aug;6(8):e006069.

7. Graham HR, Bagayana SM, Bakare AA, Olayo BO, Peterson SS, Duke T, et al. Improving Hospital Oxygen Systems for COVID-19 in Low-Resource Settings: Lessons From the Field. Glob Health Sci Pract. 2020 Dec 23;8(4):858–62.

8. La Vincente SF, Peel D, Carai S, Weber MW, Enarson P, Maganga E, et al. The functioning of oxygen concentrators in resource-limited settings: a situation assessment in two countries. Int J Tuberc Lung Dis Off J Int Union Tuberc Lung Dis. 2011 May;15(5):693–9.

9. Subhi R, Burhin M, Lam F, Webster H, Drucker E, Kamuntu Y, et al. Medical oxygen service readiness and service coverage in seven countries in Africa and Asia: a cross-sectional survey and clinical audit. Lancet Glob Health. 2025 Oct 1;13(10):e1701–14.

10. Schedwin M, Graham H, Losneanu A, Houdek J, Kitutu FE, Mukisa P, et al. Exploring the Role of ‘Outsourced Oxygen-to-the-Bedside’ in Expanding Oxygen Access for Patients. Preprints [Internet]. 2024 Dec 5; Available from: 10.20944/preprints202412.0478.v1

11. Lam F, Stegmuller A, Chou VB, Graham HR. Oxygen systems strengthening as an intervention to prevent childhood deaths due to pneumonia in low-resource settings: systematic review, meta-analysis and cost-effectiveness. BMJ Glob Health [Internet]. 2021 Dec 20 [cited 2025 May 6];6(12). Available from: https://gh.bmj.com/content/6/12/e007468

12. Graham H, Tosif S, Gray A, Qazi S, Campbell H, Peel D, et al. Providing oxygen to children in hospitals: a realist review. Bull World Health Organ. 2017 Apr 1;95(4):288– 302.

13. Smith V, Changoor A, McDonald C, Barash D, Olayo B, Adudans S, et al. A comprehensive approach to medical oxygen Ecosystem building: an implementation case study in Kenya, Rwanda, and Ethiopia. Glob Health Sci Pract [Internet]. 2022 [cited 2024 Sep 9];10(6). Available from: https://www.ghspjournal.org/content/10/6/e2100781?utm_source=TrendMD&utm_medium=cpc&utm_campaign=Global_Health%253A_Science_and_Practice_TrendMD_0

14. Smith V, Changoor A, Rummage S, Wolde HF, Zeleke EG, Belay GM, et al. An Oxygen Supply Is Not Enough: A Qualitative Analysis of a Pressure Swing Adsorption Oxygen Plant Program in Ethiopian Hospitals. Glob Health Sci Pract [Internet]. 2024 Aug 27 [cited 2025 May 6];12(4). Available from: https://www.ghspjournal.org/content/12/4/e2300515

15. Damschroder LJ, Aron DC, Keith RE, Kirsh SR, Alexander JA, Lowery JC. Fostering implementation of health services research findings into practice: a consolidated framework for advancing implementation science. Implement Sci. 2009 Aug 7;4(1):50.

16. Damschroder LJ, Reardon CM, Widerquist MAO, Lowery J. The updated Consolidated Framework for Implementation Research based on user feedback. Implement Sci. 2022 Oct 29;17(1):75.

17. Proctor E, Silmere H, Raghavan R, Hovmand P, Aarons G, Bunger A, et al. Outcomes for Implementation Research: Conceptual Distinctions, Measurement Challenges, and Research Agenda. Adm Policy Ment Health. 2011;38(2):65–76.

18. Tong A, Sainsbury P, Craig J. Consolidated criteria for reporting qualitative research (COREQ): a 32-item checklist for interviews and focus groups. Int J Qual Health Care. 2007 Dec 1;19(6):349–57.

19. About the Oxygen CoLab [Internet]. Better Futures CoLab. [cited 2025 May 6]. Available from: https://www.makingbetterfutures.org/aboutoxycolab

20. Guide to infrastructure and supplies at various levels of health care facilities Emergency and Essential Surgical Care (EESC) [Internet]. Geneva, Switzerland: World Health Organization; 2012. Available from: https://www.who.int/docs/default-source/integrated-health-services-(ihs)/csy/surgical-care/imeesc-toolkit/equipment-lists-and-needs-assessment/anaesthetic-infrastructure-supplies.pdf?sfvrsn=a2aa7580_5

21. Gale NK, Heath G, Cameron E, Rashid S, Redwood S. Using the framework method for the analysis of qualitative data in multi-disciplinary health research. BMC Med Res Methodol. 2013 Sep 18;13(1):117.

22. Graham HR, Bakare AA, Gray A, Ayede AI, Qazi S, McPake B, et al. Adoption of paediatric and neonatal pulse oximetry by 12 hospitals in Nigeria: a mixed-methods realist evaluation. BMJ Glob Health. 2018;3(3):e000812.

23. Bakare AA, Salako J, King C, Olojede OE, Bakare D, Olasupo O, et al. ‘Let him die in peace’: understanding caregiver’s refusal of medical oxygen treatment for children in Nigeria. BMJ Glob Health [Internet]. 2024 May 16 [cited 2025 Dec 12];9(5). Available from: https://gh.bmj.com/content/9/5/e014902

24. Howie SR, Ebruke BE, Gil M, Bradley B, Nyassi E, Edmonds T, et al. The development and implementation of an oxygen treatment solution for health facilities in low and middle-income countries. J Glob Health. 2020 Dec;10(2):020425.

25. Bakare AA, Graham H, Ayede AI, Peel D, Olatinwo O, Oyewole OB, et al. Providing oxygen to children and newborns: a multi-faceted technical and clinical assessment of oxygen access and oxygen use in secondary-level hospitals in southwest Nigeria. Int Health. 2020 Jan 1;12(1):60–8.

26. Smith V, Changoor A, Rummage S, Wolde HF, Zeleke EG, Belay GM, et al. An Oxygen Supply Is Not Enough: A Qualitative Analysis of a Pressure Swing Adsorption Oxygen Plant Program in Ethiopian Hospitals. Glob Health Sci Pract. 2024 Aug 27;12(4):e2300515.

27. Schedwin M, Graham H, Losneanu A, Houdek J, Kitutu F, Mukisa P, et al. Exploring the Role of ‘Outsourced Oxygen-to-the-Bedside’ in Expanding Oxygen Access for Patients. 2024.

28. Sessions KL, Ruegsegger L, Mvalo T, Kondowe D, Tsidya M, Hosseinipour MC, et al. Focus group discussions on low-flow oxygen and bubble CPAP treatments among mothers of young children in Malawi: a CPAP IMPACT substudy. BMJ Open. 2020 May;10(5):e034545.

29. Adeoti AO, Desalu OO, Elebiyo T, Aremu OA. Misconception on Oxygen Administration among Patients and Their Caregivers in Ado Ekiti, Nigeria. Ann Afr Med. 2022;21(3):269–73.

